# Socioeconomic correlates of urban mobility trends in two Australian cities during transitional periods of the COVID-19 pandemic

**DOI:** 10.1101/2024.01.31.24302105

**Authors:** Pratyush Kollepara, Subhrasankha Dey, Martin Tomko, Erika Martino, Rebecca Bentley, Michele Tizzoni, Nicholas Geard, Cameron Zachreson

## Abstract

During the COVID-19 pandemic, both government-mandated lockdowns and discretionary changes in behaviour combined to produce dramatic and abrupt changes to human mobility patterns. To understand the socioeconomic determinants of intervention compliance and discretionary behavioural responses to epidemic threats, we investigate whether changes in human mobility showed a systematic variation with socioeconomic status during two distinct periods of the COVID-19 pandemic in Australia. We analyse mobility data from two major urban centres and compare the trends during mandated stay-at-home policies and after the full relaxation of NPIs, which coincided with a large surge of COVID-19 cases. We analyse data aggregated from de-identified GPS trajectories, collated from providers of mobile phone applications and aggregated to small spatial regions. Our results demonstrate systematic decreases in mobility relative to pre-pandemic baseline with the Index of Education and Occupation, for both pandemic periods. On the other hand, the Index of Economic Resources was not correlated with mobility changes. This result contrasts with observations from other national contexts, where reductions in mobility typically increased strongly with indicators of wealth. We interpret these findings in the context of the economic policies put in place by Australian authorities to subsidise household incomes and maintain the essential workforce.

## I. INTRODUCTION

Equitable public health responses to pandemics such as COVID-19 require an understanding of the socioeconomic correlates of behaviour associated with intervention policies and epidemic dynamics [1]. Public health interventions put in place during the COVID-19 pandemic affected human mobility across all spatial scales, from the effects of international travel restrictions to the micro-distancing policies put in place to decrease crowd density in public spaces [2]. Due to the widespread use of mobile devices, changes in mobility that occurred during COVID-19 have been the subject of unprecedented levels of analysis and have proven useful for understanding the many factors influencing behaviour change during the pandemic [3–11].

Research on the relationship between changes in mobility during COVID-19 lockdown periods as a function of socioeconomic status has been conducted in many countries, using mobility data, or through self-reported activity surveys. Some studies use a single measure of socioeconomic status (SES) while others consider various components of SES such as education, occupation, and income. Studies that use a single aggregate measure of SES have reported that mobility or behaviour change is the least pronounced for those with low SES [12–14]. Studies that consider the components of SES indices have found differing trends of mobility with the SES components, such as a decrease in mobility with income [15–17], decrease in mobility with education but no variation with income [18, 19], decrease in mobility with income but no variation with education level [20] and decrease in mobility with both wealth and occupational status [21]. While such studies demonstrate that socioeconomic conditions are important considerations for understanding and predicting behavioural responses to interventions, the lack of consensus illustrates the contextual complexity of these questions. Our study aims to analyse complex drivers of movement behaviour by comparing periods exhibiting contrasting behavioural stimuli.

In this study, we investigate mobility changes within Australian urban centres during two phases of the pandemic. We aim to understand the differences between the behavioural response to mandated stay-at-home policies, implemented during the early phases (April 2020), and the response to the full relaxation of those policies which occurred almost two years later (January 2022). Our study leverages de-identified GPS data, collated from providers of mobile phone applications and aggregated to small spatial regions by the Australian data analytics company *Pathzz*. By assessing changes to mobility levels from each region as a function of socioeconomic characteristics (assessed by the Australian Census) we identify socioeconomic drivers of mobility behaviour change during two distinct periods of the COVID-19 pandemic in Australia.

Our investigation focuses on the two largest Australian cities (Sydney and Melbourne), and we examine two scenarios: the initial period of stay-at-home orders, and the re-opening phase that coincided with a wave of infections caused by the Omicron variant. Mandated stay-at-home policies were introduced at the end of March, 2020 (Figure 1). During this period disease prevalence was low, risk perception was almost universally high, and stay-at-home orders were supported by the Australian Government both through financial support packages and through large financial penalties introduced to enforce the measures [22, 23]. In the second scenario (re-opening), stay-at-home orders were universally relaxed, along with all other travel restriction policies introduced throughout the pandemic. Though stay-at-home orders were not in effect, some prevention policies were in place (scanning QR codes for contact tracing, mask wearing in certain spaces etc.) [22, 24, 25]. During this period, risk perception remained high for large sections of the population. Policy relaxation, combined with the emergence of the newly dominant Omicron variant, produced a surge in confirmed COVID-19 cases in January of 2022. This surge in cases resulted in what was referred to as “shadow lockdown” i.e., substantial discretionary reductions in movement and social activity resulting from the perceived risk of infection [26]. [Note: the term “shadow lockdown” should not be confused with the similar term “shadow pandemic” which is used in other work regarding domestic violence during COVID-19 [27]].

**FIG. 1.**
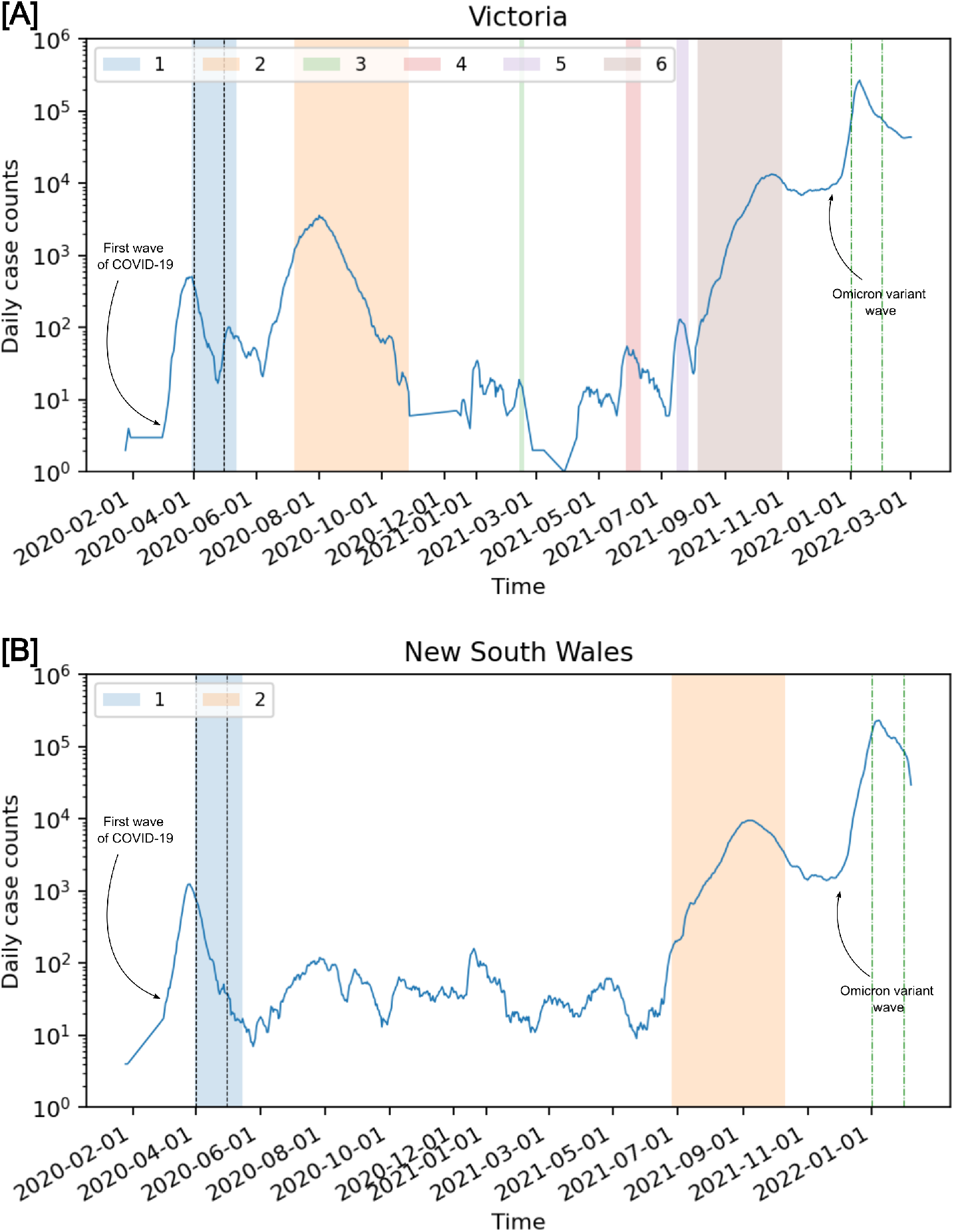
A timeline of COVID-19 cases and stay-at-home policies in the states of Victoria (a) and New South Wales (b) in Australia [29, 30]. The blue line plot shows daily new cases (with a 7 day rolling average). The colored patches show the various intervention periods. The two pairs of vertical lines show the periods for which we analysed mobility trends: April 2020 and January 2022. The timelines of stay-at-home policies were obtained from an Australian parliamentary report, news articles and media releases from state governments [22, 24, 25, 31–34].

To characterise socioeconomic variation in Sydney and Melbourne, we use two aggregate indices of socioeconomic status, which characterise broadly distinct categories of factors. Specifically, one of the indices exclusively quantifies economic resources (ER) while the other combines education and occupational factors (EO). While highly correlated (Figure 2), these indices are formed from separate sets of component variables measured by the Australian Census [28].

**FIG. 2.**
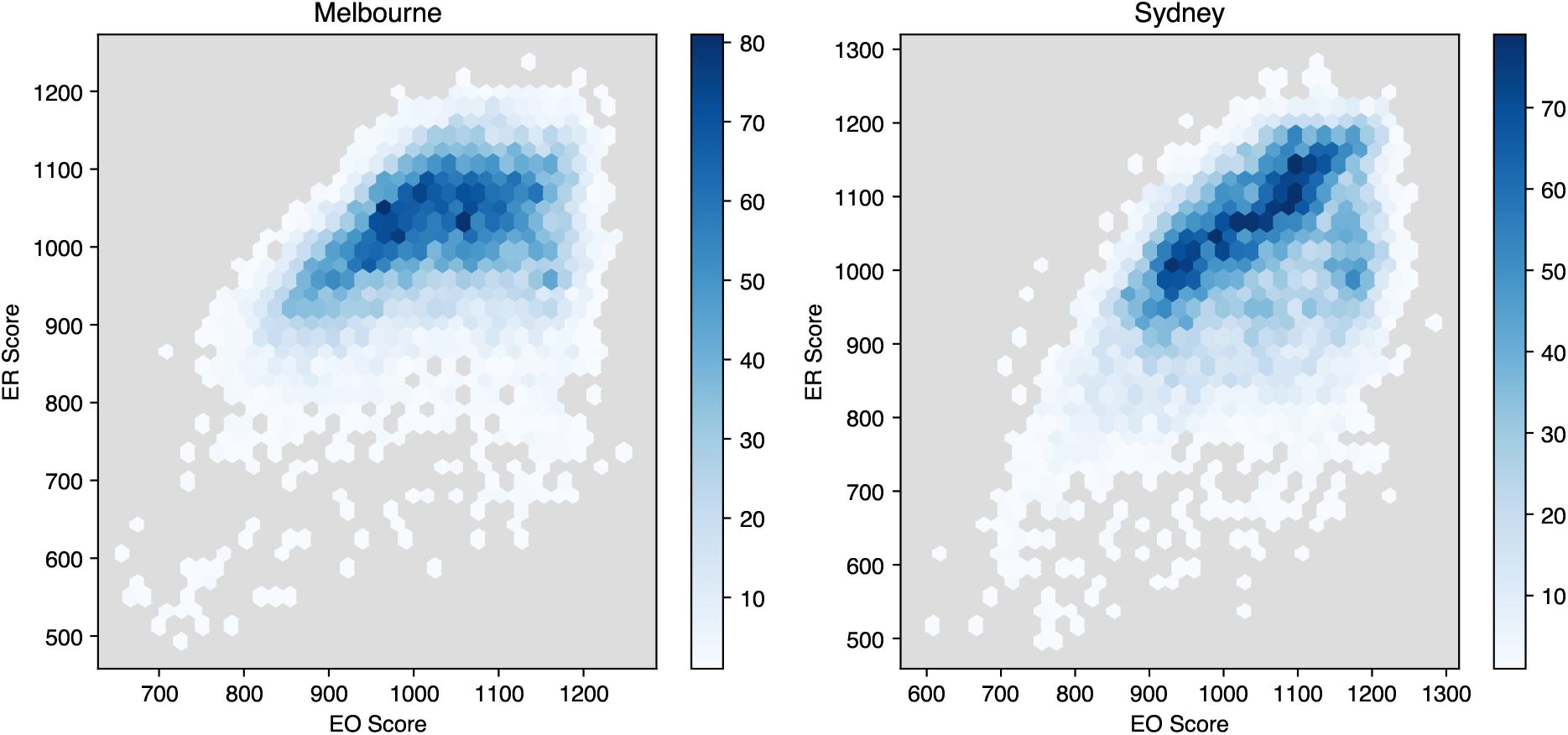
Joint distribution of Index of Education and Occupation (EO) and Index of Economic Resources (ER) scores in Melbourne and Sydney.

Our findings demonstrate that, for both periods, economic resources were not associated with differences in mobility behaviour. One the other hand, education and occupation factors were associated with reduced mobility during both the initial period of stay-at-home orders, and the re-opening phase (coinciding with the Omicron wave). Notably, these differences in mobility behaviour with socioeconomic indicators occur despite the fact that EO and ER indices are highly correlated, requiring an analysis of mobility across their joint distribution. This joint analysis demonstrates that the effects of ER competed with those of EO, producing a trend in which regions with high EO index, but low ER index exhibited lower levels of mobility compared to those with low EO and high ER.

## II. METHODS

### A. Scenario Descriptions

#### 1. Pandemic conditions in April 2020 and January 2022

Before the introduction of stay-at-home orders, daily counts of confirmed COVID-19 infection increased through most of March 2020, with the trend reversing by the end of March in both Victoria and New South Wales and more than 500 and 1000 daily cases reported at peak levels in the two states. This reversal coincided with stay-at-home orders and international travel restrictions. By the end of March 2020, a total of 2153 cases had been detected in NSW and 956 cases had been detected in Victoria. During April 2020, the first period we analyse in our study, case counts broadly declined in both states. By the end of April, 3018 cases had been detected in NSW and 1365 in Victoria. During the second period of interest, January 2022, daily case counts were in the order of 10^5^ in both states, due to the recent introduction of the Omicron variant and relaxation of restrictions [35].

#### 2. Policy conditions in April 2020 and January 2022

In Victoria, activity restrictions to combat transmission of COVID-19 had increased in intensity over time to include: banning non-essential gatherings of size more than 500 persons, banning indoor gatherings of more than 100 persons, restrictions on visits of aged care homes, and closure of non-essential services. On the 31st of March 2020, “stage three” restrictions were announced. Only four reasons were prescribed as a justification for an individual to leave their home: to acquire food and supplies, to access healthcare, to exercise, and for the purpose of work/education. Gatherings of more than 2 persons were not allowed, unless the individuals belonged to the same household and for work/education. Playgrounds, skateparks and outdoor gyms were closed. On-the-spot fines of AUD$ 1652 (worth minimum wage work of 71 hours) for individuals and AUD$ 9913 for businesses were enforced if any of these restrictions were breached.

All schools transitioned to remote and flexible teaching [22].

In New South Wales, activity restrictions included: banning non-essential indoor gatherings of more than 100 persons, and outdoor gatherings of more than 500 persons, restrictions on visits to aged care homes, enforced social distancing of 1.5 metres, and closure of non-essential businesses and activities. On the 31st of March 2020, New South Wales issued a set of valid reasons for leaving one’s home, as did Victoria [22, 36]. Relief measures during this time included a national moratorium on evictions and monetary payments to more than 5 million people, among other measures and supports for housing [22, 23].

Between April 2020 and December 2021, Victoria had gone through 5 subsequent lockdown periods of varying intensity and duration, while New South Wales had one subsequent lockdown. By January 2022, major restrictions on movement and gatherings were removed across the country (conditional on vaccination), though density limits and mask mandates remained [31–34].

#### 3. Baseline periods

The baseline periods used for comparing the mobility during the periods of interest were: 15 September - 15 October 2019 (for comparison with April 2020, the lockdown period) and January 2020 (for comparison with January 2022, the omicron wave period) (Table I). Annually-matched mobility data from April 2019 was not available for comparison with April 2020, so we selected the period between September 15 and October 15 because it has a similar number of school holiday dates which we expect to influence mobility patterns. In January 2020, Australian states had detected their first COVID-19 cases, but interventions were not in place other than screening procedures at international airports [22].

**TABLE I.** Periods of interest and the corresponding pre-pandemic baseline periods used for comparison.

| Description | Test period | Baseline period |
| --- | --- | --- |
| Lockdown period | April 2020 | 15th September - 15th October 2019 |
| Omicron wave period | January 2022 | January 2020 |

### B. Data source

Mobility data gathered from mobile device GPS traces was provided for the purposes of this study by the data analytics company Pathzz (https://www.pathzz.com/). Pathzz is a consumer intelligence platform that uses mobility signals from opted-in smartphone devices in order to provide movement analytics, quantifying how people interact with spaces. The data is used by government organisations, private businesses, investment banks and research firms. For comparison with local population characteristics, users are assigned a statistical area as a home location [Statistical Area Level 1 (SA1), Australian Statistical Geography Standard [37]]. Home locations are assigned to devices based on where they are located most often between the hours of 6pm to 8am Monday through Friday, and for the entire weekend period (Saturday and Sunday). By aggregating to home locations, mobility trends can be analysed with respect to aggregate statistical data from the Australian census and other sources. In Australia, Pathzz data is derived from approximately 17M unique devices per year, though the number of devices accounted for at at given time varies due to fluctuating user numbers, use of multiple devices by individuals, and device upgrades.

Using the Pathzz platform, a user begins by defining the destinations and time periods of interest. The Pathzz platform then provides the number of visits by home location, over all SA1 regions within 50km of the specified destination. Visits are counted when a unique device enters the user-specified geospatial boundary during the specified period, and a maximum of one visit per calendar day is recorded for each device.

Compared to alternate sources of aggregate mobility data, Pathzz provides a flexible and high-resolution database. The capacity to define customised sets of destination locations allowed us to apply our analysis to entire urban regions, at high spatial resolution. Simultaneously, Pathzz protects the privacy of individual devices by restricting analysis to aggregate home locations. Pathzz does not provide individual device trajectories or link visits to personal information associated with devices (other than putative home regions) which helped us manage the risk of inadvertently violating user privacy during our study. Due to the spatial heterogeneity of socioeconomic status, and the high spatial resolution of the data, our study benefited from increased variance of socioeconomic status between regions which would otherwise be lost due to spatial aggregation.

### C. Indices of Economic Resources (ER) and Education & Occupation (EO)

The Australian Bureau of Statistics publishes a set of four indices referred to as Socio-Economic Indexes for Areas (SEIFA) that quantifies the relative social and economic advantages and disadvantages of statistical regions at various spatial scales [28]. The indices are: Relative Socio-economic Advantage and Disadvantage (IRSAD), Index of Relative Socio-economic Disadvantage (IRSD), Index of Education and Occupation (IEO), Index of Economic Resources (IER). Both IRSAD and IRSD reflect advantage and disadvantage in terms of combined social and economic factors. On the other hand, the indices of EO and ER comprise of variables that reflect only social advantage/disadvantage and economic advantage/disadvantage, respectively. ER and EO indices do not have any common component variables. Because ER and EO indices examine non-overlapping sets of factors, we chose them for our study of mobility trends by SES.

Specifically, the component factors of EO relate to educational qualifications and skill level of occupation, while those of ER are income, rental expenses and home ownership. A region will have a low EO score if: (i) many individuals are without educational qualifications, employed in low skilled occupation, or unemployment is high; and (ii) few individuals are highly qualified or employed in highly skilled occupations. A region will have a low ER score if there are: (i) many households that have low income or pay low amounts of rent; and (ii) few households with high income or few people who own their home [28]. For the analysis and results presented in this study, we stratify the ER and EO scores for the SA1 regions of each city separately, into local decile bands.

#### 1. Analysis of mobility data from Pathzz

For aggregation and processing of mobility data, space was divided into two different sets of partitions for origin locations (SA1 partitions) and destinations [Destination Zone (DZN) partitions, Australian Statistical Geography Standard]. SA1 regions contain resident populations between 200 and 800 people, with a mean of approximately 400 (Figure S2). The spatial extent of SA1 and DZN regions depends on population density, and is typically much greater on the urban fringe than in the central urban regions (Figure 3). DZN regions spatially partition workplace locations and were originally designed for use in the analysis of place-of-work surveys included in the Australian Census. These partition schemes are described by the Australian Bureau of Statistics Australian Statistical Geography Standard (ABS ASGS) [37].

**FIG. 3.**
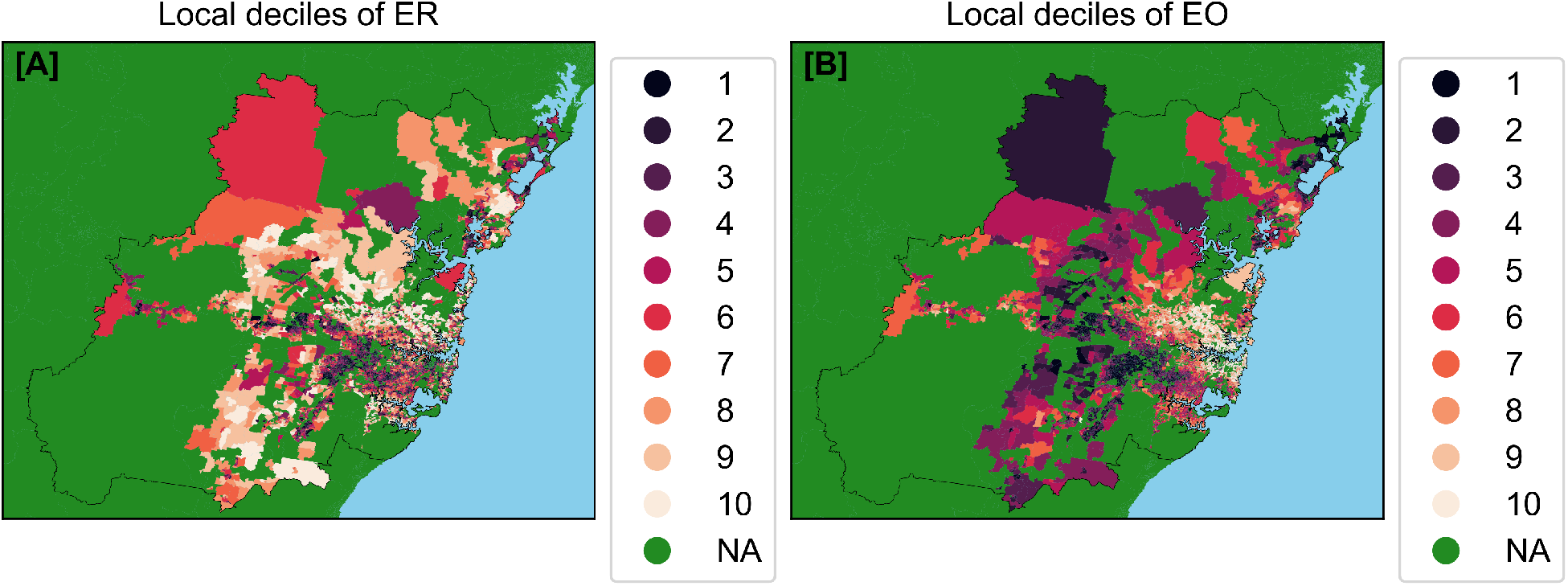
Choropleth maps of the Greater Sydney urban region with colors corresponding to insample decile scores for socioeconomic indices of Economic Resources (ER) [A] and of Education and Occupation (EO) [B]. Dark colors correspond to areas with lower SES and green regions were not included in the analysis. The thin black line represents the boundary of the greater urban region. Note that while ER and EO are strongly correlated, areas with high EO and low ER tend to be located closer to the interior of the city, while regions with high ER and low EO tend to be located towards the urban fringe. Similar spatial trends are observable in the Greater Melbourne area (see supporting information Figure S4).

For each DZN partition, data was collected from the Pathzz platform by specifying a time period (corresponding to the test periods and respective baseline periods) and the unique spatial polygon describing the DZN of interest. After specifying the DZN geometry, visitation counts from all SA1 regions were downloaded from Pathzz in the format of a “Data Analytics Report”, which provides aggregate counts of all visits into the specified region that originated from each SA1 partition with a 50km radius, over the specified time period. This count data from the entire set of DZN regions within Greater Sydney and Greater Melbourne was then disaggregated by origin to generate a full origin-destination matrix structured as a directed edge list (SA1 → DZN). Finally, these pair-wise counts were re-aggregated as mean counts per day by origin.

Within a DZN region, a visit of any duration is recorded and counted with equal weight. For this reason, adjacent and intersecting SA1 and DZN partition pairs were excluded from our analysis of mobility trends (Figure 4a). Unlike other sources of mobility data (such as that provided by Meta’s data-for-good program), which are temporally filtered and can be interpreted as origin-destination matrices, the data described here should be interpreted as “pass-through” counts (i.e., a visit should not be interpreted as a trip from an origin region and an individual’s final destination). If an individual passes through multiple DZN regions during a single day, they will be counted once in each region through which they pass. For this reason, we restrict our analysis to the comparison of visit counts, aggregated by origin SA1, relative to baseline (i.e., we implicitly assume that the rate at which origin-destination journeys are double-counted depends on the origin, but not on the time period over which data is aggregated).

**FIG. 4.**
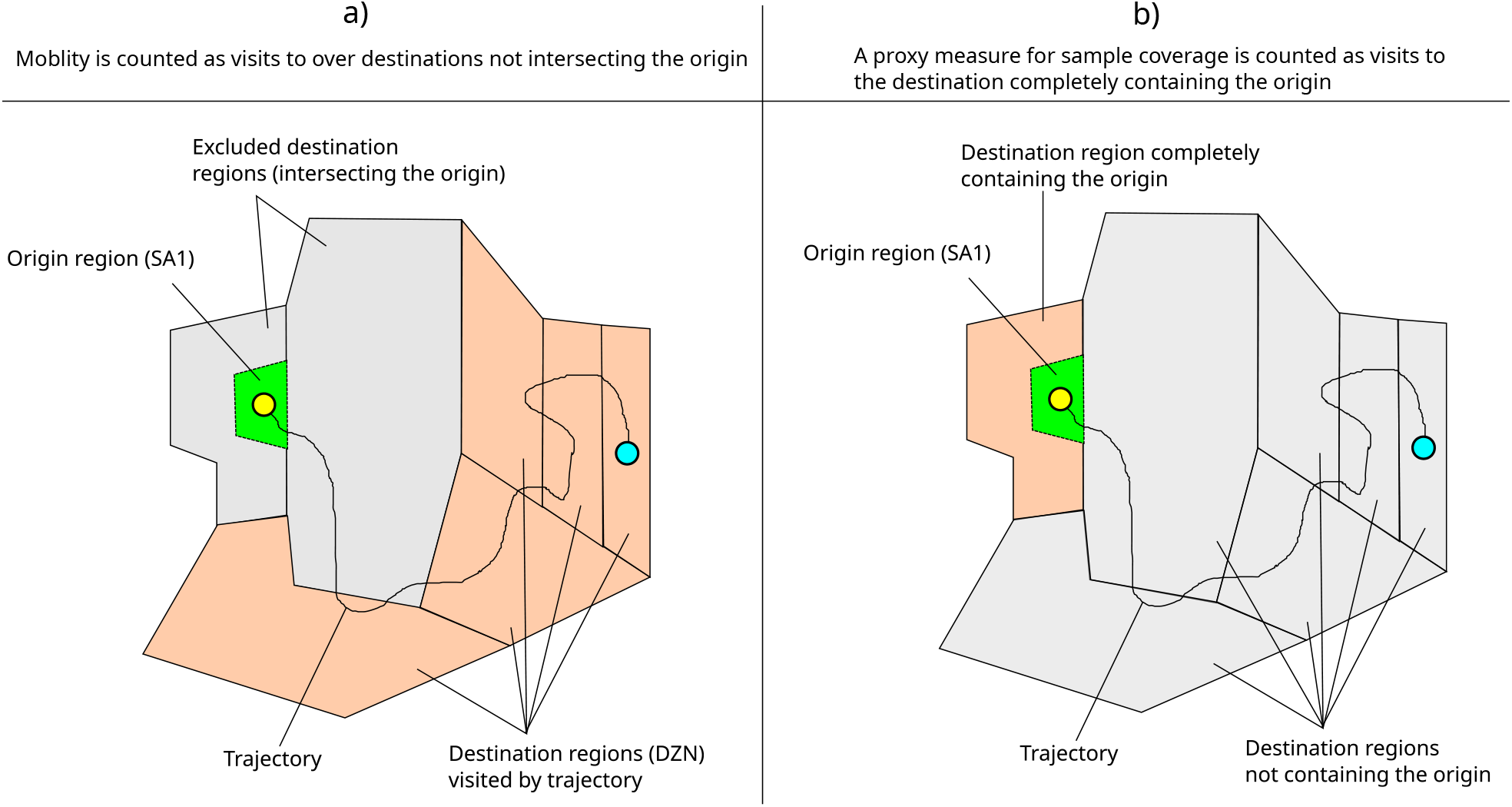
Schematic representations of mobility data aggregation for: (a) the method for computing counts of visits between origin regions and destination regions. A single individual trajectory is represented for illustrative purposes. Visits counted between the origin SA1 region and destination zones correspond to the number of trajectory points located within each DZN. The set of DZN regions intersecting the origin SA1 region are excluded from the count to avoid including spurious visits due to GPS jitter and local movement within origin regions. (b) Sample coverage is computed as the number of trajectory visits counted within the destination area completely containing the origin region.

For each origin region and time period of interest, we assess changes in mobility as the log-transformed ratio of trip counts during test and baseline periods, expressed as proportions of sample coverage over each time window:

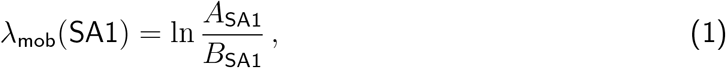

where

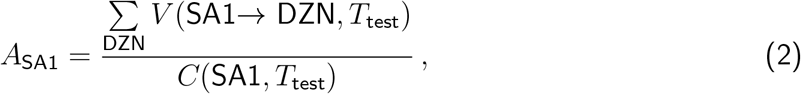

is the number of visits *V* recorded for an SA1 origin region, summed over all non-intersecting destinations and averaged per day over the duration Δ*t*_test_ of the period of interest *T*_*test*_ (either after introduction of stay-at-home orders, or during re-opening), divided by the sample coverage *C* recorded over the same period and

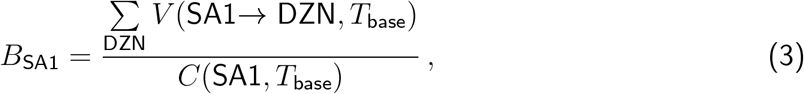

is the same measure for the corresponding pre-pandemic baseline interval *T*_base_.

#### 2. External validation

To examine whether or not the data acquired from Pathzz was consistent with independent measures of mobility trends, we compared it to intersection pass-through data available through the Victorian Department of Transportation. This external validation is described in full in the Supporting Information. Robust positive correlations were found for all periods, indicating that the mobility data acquired from Pathzz is generally consistent with other sources of mobility data.

## III. RESULTS

### A. Overall mobility trends

Following the introduction of stay-at-home orders, median mobility decreased substantially in both urban regions, as shown by histograms of λ_mob_ over SA1 partitions (Figure 5). These distributions also show high variability in λ_mob_, demonstrating that while mobility decreased overall, large numbers of SA1 regions exhibited higher mobility during lockdown, relative to baseline.

**FIG. 5.**
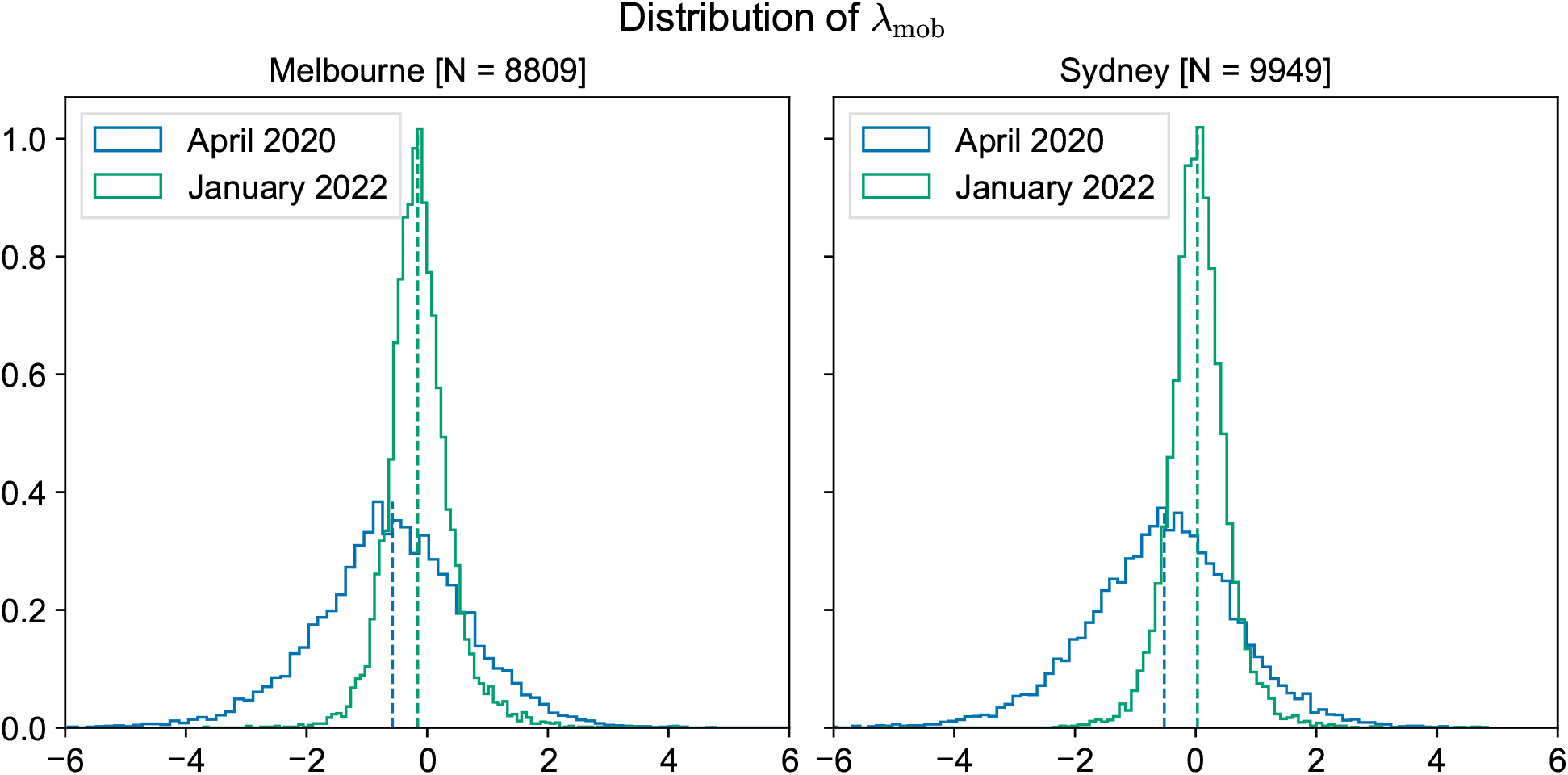
Histograms over SA1 regions of *λ*_mob_, the log-transformed ratio of mobility levels during COVID-19 periods and baseline periods. Blue traces correspond to the initial period of stay-at-home orders in April 2020, and green traces correspond to the Omicron surge and re-opening period in Jan 2022. The vertical dashed lines show the median value of the distribution with the corresponding color.

After the relaxation of mobility restrictions, international travel bans, and regulations on social gatherings (the re-opening phase), there was a large wave of COVID-19 cases attributed to the Omicron variant. Overall changes in mobility are shown as distributions over SA1 partitions (Figure 5), which shows a slight median decrease in Melbourne, and no significant change in median mobility in Sydney. Distributions for both regions demonstrate substantial variability, with approximately even proportions of SA1 regions showing higher or lower mobility relative to baseline.

### B. Mobility as a function of socioeconomic indices

In both Melbourne and Sydney, λ_mob_ declines with EO, but does not change with ER during the period of stay-at-home orders in April 2020, with small but significant correlations between EO scores and λ_mob_ values for both urban regions (Figure S5). To more clearly illustrate these trends, we stratify the SA1 regions into local decile bands for EO and ER and plot decile medians (Figure 6 and Figure 7). We quantify the robustness of trends in decile medians, by assessing their monotonicity with decile rank [Mann-Kendall test, Table II] [38, 39]. This analysis demonstrates visible downward trends and significant monotonicity of decile medians of λ_mob_ (Figure 6a,b, Table II).

**FIG. 6.**
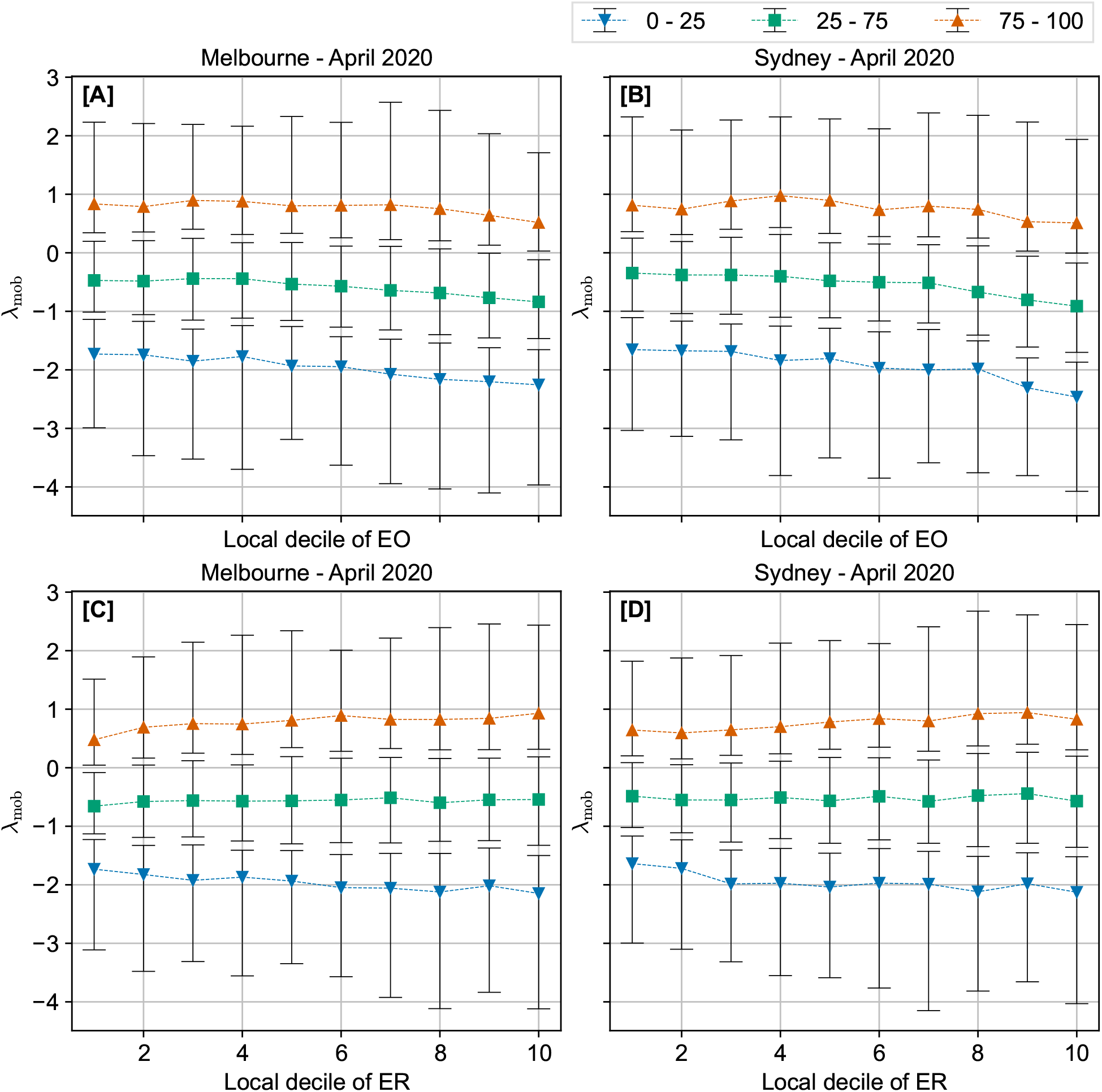
Mobility trends during stay-at-home orders as a function of socioeconomic indices. Summary statistics of *λ*_mob_, the log-transformed ratio of mobility relative to baseline are shown as a function of Education and Occupation index (EO) (a,b), and as a function of Economic Resources index (ER) (c, d). Marked line plots demonstrate summary statistics over SA1 partitions of Australian cities of Melbourne (a, c) and Sydney (b, d) during the month of April 2020. The down-facing triangles represent the median of the first quartile, squares represent the median, while the up-facing triangles represent the median of points above the third quartile. The error bars represent the 5th and 95th percentiles of the data.

**FIG. 7.**
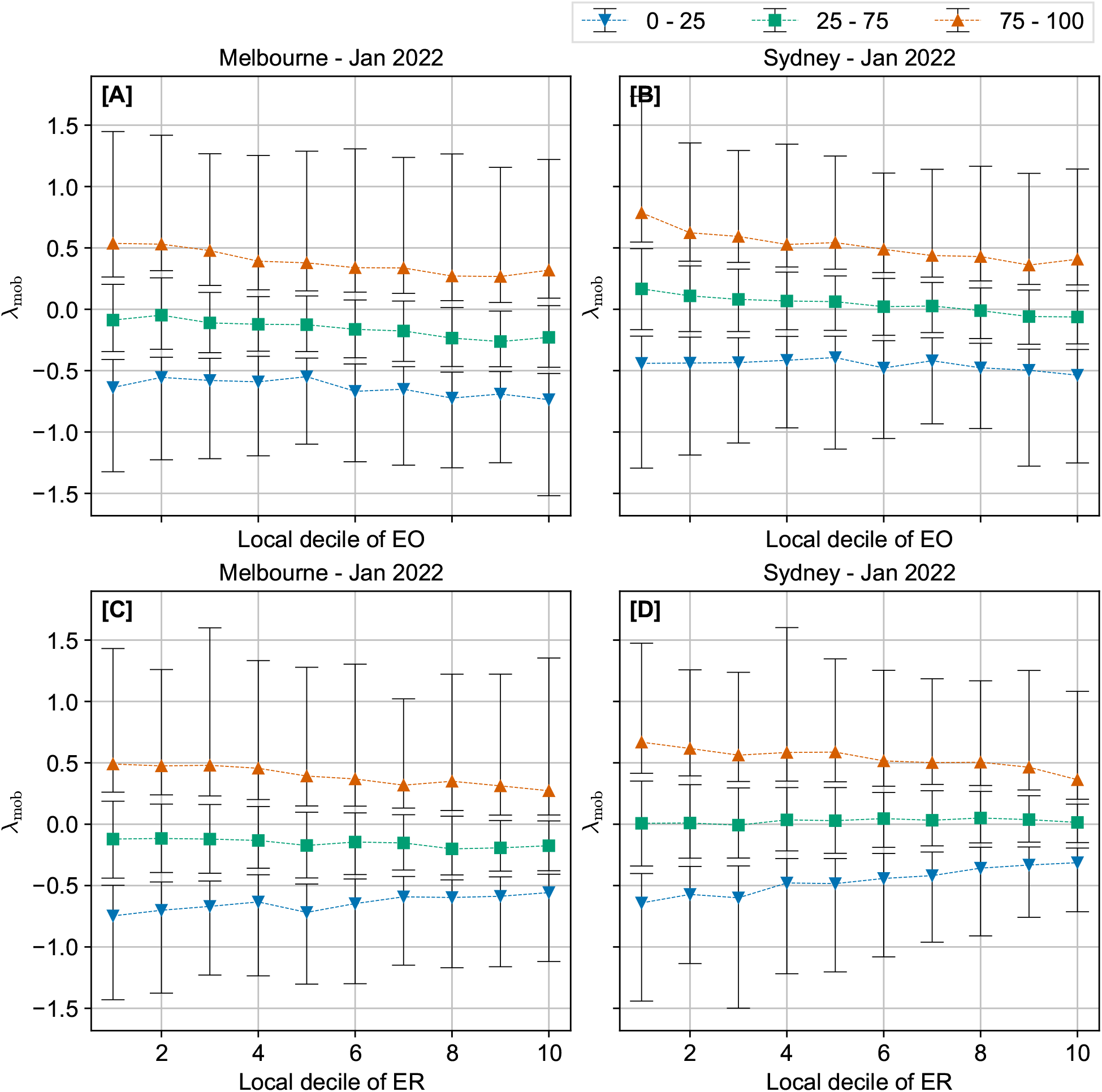
Mobility trends during the re-opening phase (and Omicron wave) as a function of socioeconomic indices. Summary statistics of *λ*_mob_, the log-transformed ratio of mobility relative to baseline are shown as a function of Education and Occupation index (EO) (a,b), and as a function of Economic Resources index (ER) (c, d). Marked line plots demonstrate summary statistics over SA1 partitions of Australian cities of Melbourne (a, c) and Sydney (b, d) during the month of January 2022. The down-facing triangles represent the median of the first quartile, squares represent the median, while the up-facing triangles represent the median of points above the third quartile. The error bars represent the 5th and 95th percentiles of the data.

**TABLE II.** Mann-Kendall test results between SEIFA deciles and the median values of *λ*_mob_. Yellow cells correspond to significant correlations between SEIFA scores and *λ*_mob_ (See the supporting information, Figure S5 and Figure S6). For ten data points, the Mann Kendall score can take odd integer values between -45 (monotonically decreasing) and +45 (monotonically increasing).

|  | Region | Greater Melbourne |  | Greater Sydney |  |
| --- | --- | --- | --- | --- | --- |
| | Mann-Kendall | score | $p$-value | score | $p$-value |
| Period | SEIFA |  |  |  |  |
| April 2020 | Local decile of EO | -37 | $1.28 \times 10^{-3}$ | -45 | $8.30 \times 10^{-5}$ |
| | Local decile of ER | 25 | $3.18 \times 10^{-2}$ | -1 | 1.00 |
| Jan 2022 | Local decile of EO | -39 | $6.77 \times 10^{-4}$ | -43 | $1.72 \times 10^{-4}$ |
| | Local decile of ER | -33 | $4.21 \times 10^{-3}$ | 19 | $1.07 \times 10^{-1}$ |

In Melbourne, while there is no visible trend in *λ*_mob_ as a function of ER, the monotonicity result reveals an increasing trend, significant at *p* = 0.032 (Table II), but this does not correspond to any correlation in ER score with λ_mob_ (Figure S5), so we disregard it as a numerical artefact. In Sydney we find no trend in λ_mob_ as a function of ER (with no visible trend and no monotonic relationship with decile medians).

During the reopening phase in January 2022, while overall reductions in mobility are much less pronounced than during the initial phase of restrictions, the decline of λ_mob_ with EO decile remains observable (Figure 7a,b). Likewise, no clear trend is visible between λ_mob_ and ER, for either urban region (Figure 7c,d). However, the MK test of monotonicity between decile medians of λ_mob_ and decile ranks reveals a slightly decreasing trend in Melbourne that is not observed in Sydney where a slight, but non-significant increase is observed (Table II). The trend in Melbourne reflects a weak and non-significant correlation between ER scores and λ_mob_. In Sydney, however, we see a significant but weakly increasing correlation between ER scores and λ_mob_.

The general observation, visible both periods, that trends in mobility are not the same with ER and EO conflicts with the observation that ER and EO are themselves highly correlated (Figure 2). The deviations from the expected behaviour indicates that a joint analysis of mobility over both ER and EO strata is necessary to understand the role of socioeconomic factors in mobility changes. To assess joint trends of λ_mob_ with ER and EO indices, Figure 8 shows heatmaps of median λ_mob_ values for all SA1 regions jointly stratified by EO and ER decile. The joint trends of mobility with EO and ER differ between the two urban regions. In Melbourne, while λ_mob_ tends to decrease with EO within ER deciles, there are no clear relationships between λ_mob_ and ER within EO deciles for either period (Figure 8a,c). For Sydney, both periods demonstrate jointly monotonic trends: λ_mob_ decreases for higher EO within ER deciles, and increases for higher ER within EO deciles (Figure 8b,d). The joint trend is most apparent in Figure 8d. The distribution of SA1 regions over the joint ER and EO decile bands is shown in the Supporting Information Figure S3.

**FIG. 8.**
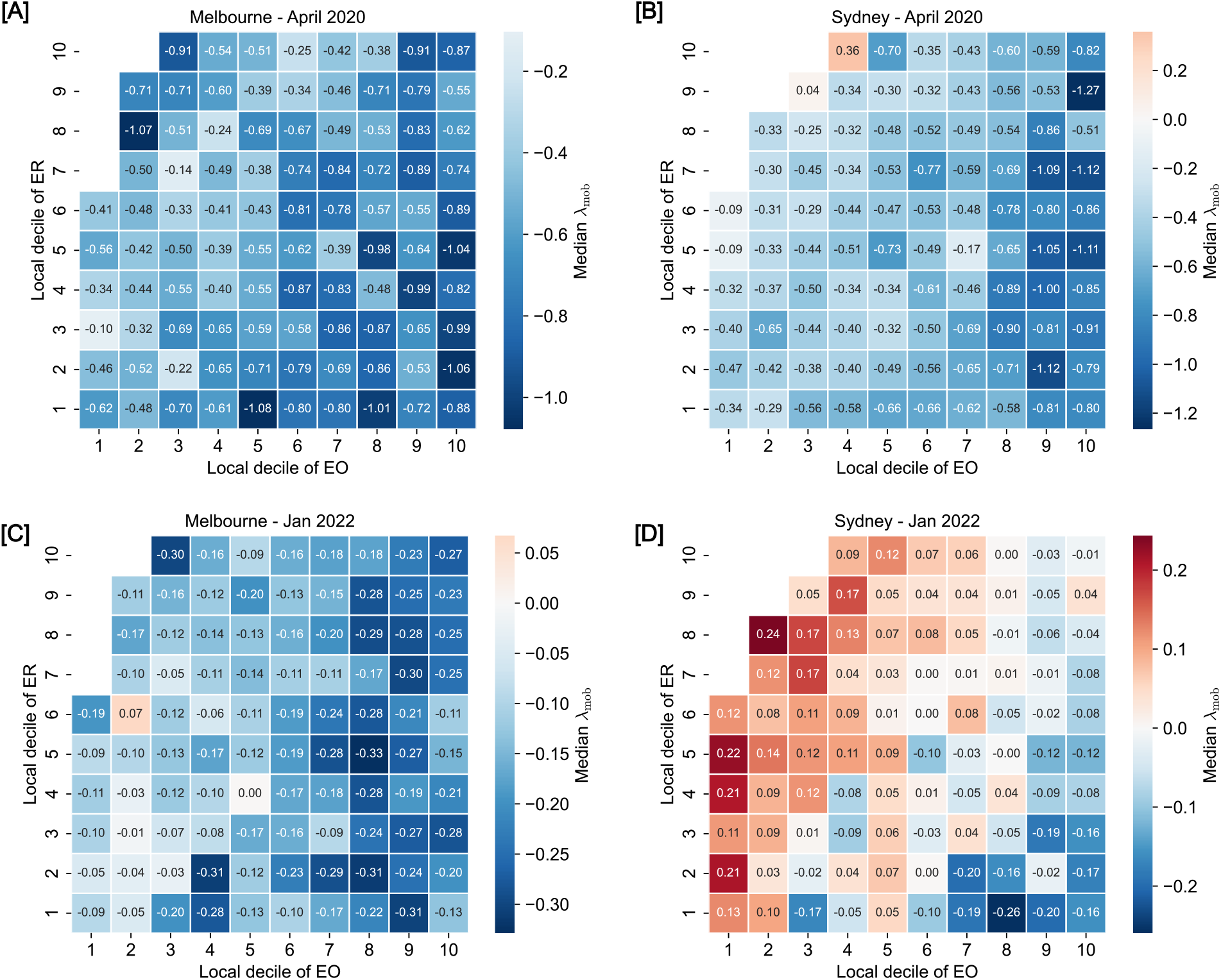
Heatmaps of median *λ*_mob_ (log-transformed ratio of mobility relative to baseline), jointly stratified by deciles of the Index of Economic Resources (ER) and Index of Education and Occupation (EO). Mobility response during the initial period of stay-at-home orders is illustrated for Melbourne (a) and Sydney (b), and the re-opening phase in Melbourne (c) and Sydney (d). Red shades correspond to increased mobility relative to baseline, light shades correspond to near-baseline mobility, and blue shades correspond to decreased mobility.

## IV. DISCUSSION

We examined trends of mobility behaviour during two turning points of the COVID-19 pandemic in Australia as functions of two different indices of socioeconomic status. During the initial period of stay-at-home orders during April 2020 we found that mobility decreased substantially overall and declined as a function of the Index of Education & Occupation, but found no relationship between mobility and the Index of Economic Resources. Surprisingly, while overall reductions in mobility were much less substantial during the re-opening phase (and Omicron wave) in January 2022, mobility trends as a function of socioeconomic indicators were similar, decreasing for higher indices of Education & Occupation, but stable as a function of Economic Resources. In Sydney, we found a prominent joint dependence of mobility on both socioeconomic indicators, producing a scenario in which regions with high ER, but low EO exhibited the highest mobility levels relative to pre-pandemic baseline.

To interpret these findings, we consider them from the frame of reference provided by the COM-B model of behaviour, which relates capability (C), Opportunity (O), and Motivation (M), to individual behaviour (B) [40]. Capability refers to attributes of individuals that make behaviours possible in principle. These could include having the sufficient mental capacity (e.g., understanding and memory) and physical capacity (e.g., physical hardware or physiological ability) to carry out the behaviour. Opportunity refers to environmental factors that facilitate the behaviour, such as proximity to locations where the behaviour can be performed, or access to the financial resources required to undertake actions necessary for the behaviour. Motivation refers to the desire to change behaviour, which can be influenced by observations and information. Motivation is mediated by opportunity and capability in the sense that if a behaviour is perceived as difficult because the conditions facilitating it are not present, the desire to do it may decrease. Finally, behaviour refers to the activities of individuals, and behaviour change occurs when an individual switches from one set of activities to another. The COM-B model emphasizes the effects of feedback between behaviours enacted, and the COM factors affecting the tendency to change behaviour.

With respect to the COM-B model, the socioeconomic indices we analysed can be considered as components of capability because they describe characteristics of individuals that mediate behaviour but are stable over long timescales. Situational assessment can provide insight into the opportunity and motivation components. Opportunity to change behaviour can be facilitated through support policies such as the economic support packages provided by the Australian Government during the initial period of stay-at-home orders, and we credit these with the notable lack of correlation between ER and mobility changes. We observed that all income strata demonstrated similarly large decreases in mobility during this crucial period, in contrast to findings from other contexts [15–17].

On the other hand, it appears the capability factors associated with education level and occupation status were associated with decreased mobility, which was observed for both locations during the initial restrictions and Omicron surge. This result is rather unique - while other studies have observed correlations between drops in mobility and education level, these past studies investigated scenarios in which mobility restrictions and COVID-19 transmission were simultaneously present [18, 19]. In contrast, our study examines a period during which mobility restrictions were absent, and the drivers of mobility behaviour change are more likely attributable to discretionary responses to epidemic prevalence and the perception of risk. This result may reflect that people with higher education level were more capable of staying informed about the risks associated with infection, helping to motivate risk avoidance behaviour. The role of occupational status is likely related to the higher proportion of people with high-skilled occupations who are able to work from home while maintaining secure employment conditions [41]. In addition, exemptions to stay-at-home restrictions were given to maintain the workforce in essential industries, many of which employ low-skilled occupations and are associated with lower EO indices (with some exceptions, such as healthcare workers) [42].

There are several limitations to our analysis that must be taken into consideration and warrant future work to validate these findings with analysis of additional data. Namely, because the data we analysed was not curated specifically for the purpose of this investigation, three methodological limitations arose that could not be fully accounted for:

1. The trajectory data underlying our analysis was not subject to temporal filtering and aggregation.

2. Sample coverage could only be computed for a subset of regions.

3. Home locations in mobility data may not correspond to true residence location.

First, because the trajectory samples were not temporally filtered, the counts of visits we analysed do not represent easily-interpreted quantifiers of origin-destination travel behaviour. In our analysis, individuals are counted multiple times as they traverse the urban landscape (once per day, for each DZN they visit), and interpretation of our observations rests on the assumption that the rate of double-counting did not change between the baseline periods and test periods analysed in our study. Second, because calculation of sample coverage required an SA1 to be completely contained by a single DZN, it could only be computed for a subset of regions. Many of the excluded regions were located in the urban fringe where SA1 and DZN geometries tend to be more extensive, with complex boundaries. Further work should account for sample coverage from all regions so that exclusions can be minimised. Finally, because a data-driven approach was used to classify the residential locations associated with de-identified device trajectories, there is no guarantee that the home location is a true representation of the user’s residential address. For this reason, the association between socioeconomic status of residential partitions (SA1 regions) and home locations of tracked devices is subject to unknown bias.

## V. CONCLUSION

Future implementation of equitable public health responses to pandemics such as COVID-19 require a better understanding of how socioeconomic factors influence behaviour. In this study, we found that mobility decreased with the Index of Education & Occupation for small spatial partitions of the urban landscape in the cities of Melbourne and Sydney, Australia. This correlation was observed both during stay-at-home orders, when case incidence was low, and during the reopening phase, when case incidence was peaking and restrictions were not in place. In contrast to observations in other national contexts, wealth (as quantified by the Index of Economic Resources) was not found to be correlated with mobility trends for either period. As the Indices for Economic Resources and Education & Opportunity are generally highly correlated, these contrasting trends indicate opposing effects by each Index. We tentatively conclude that these observations reflect three underlying causes: 1) the availability of economic support to facilitate compliance with stay-at-home orders; 2) the spatial clustering of essential workers who remained mobile during the initial period of restrictions and 3) the capacity of those with highly skilled employment to work from home. Additionally, we speculate that while the combined effects of low economic resources and high education level led to risk-averse behaviour during the re-opening phase, the opposite was true for those with high economic resources and low education level. These findings can help facilitate the design of equitable and effective pandemic interventions in future scenarios by identifying factors which can be used to help predict the heterogeneous behavioural responses to policy implementation and relaxation.

## VI. ETHICS STATEMENT

This project was approved by the University of Melbourne Office of Research Ethics and Integrity, application number 2022-24646-29913-3. The Committee agreed to approve the application on the basis that it met the requirements of the National Statement on Ethical Conduct in Human Research [43].

## VII. DATA AVAILABILITY

All data and code necessary for reproducing our main analysis is provided in the linked repository. This includes aggregate mobility counts and coverage levels for all SA1 regions for all time periods analysed. We have not released raw data downloaded from the Pathzz platform.

Repository: https://github.com/praty-k/Aus-ses-mob

## VIII. ACKNOWLEDGEMENTS

We would like to thank Jason Sandiego at Pathzz for offering access to their database and facilitating consultation regarding the data collection methods.

## Supporting Information

### S1. DETAILED METHODS

#### A. Exclusion criteria

Of the entire set of SA1 areas within 50km of the Greater Urban regions of Sydney and Melbourne, the following factors contributed to exclusion of regions from the set of areas we analysed in our study:

- SA1 regions for which one of the attributes [Usual Resident Population (URP), ER score and EO score] were not present or if either the base period counts or the test period counts are zero.
- Coverage could not be computed for SA1 regions that were not completely contained by a single DZN region, excluding them from the analysis. As a result, 1557 and 1077 SA1 regions were excluded from Greater Melbourne and Greater Sydney, respectively.

The exclusion statistics are summarised in Table S1. Calculation of coverage, which controls for fluctuating sample populations, is the primary contributor of data loss.

#### B. External validation: comparison with intersection passthrough counts

To validate the data provided by Pathzz against independent measurements of mobility trends, we compared the trip counts aggregated for each DZN to intersection traffic counts from the same areas. Note that this aggregation of the Pathzz data differs from that used in our main analysis. Here, we compare the total number of visits to each DZN with an independent measurement of traffic through road intersections within the same areas. While intersection traffic counts are not an exact indicator of all passthrough traffic for each DZN, we expect to find positive correlation between these measures. Furthermore, we expect to find positive correlation when we compare changes in count numbers between test and baseline periods for each measure.

**TABLE S1.** Sample sizes (number of SA1 regions) for Greater Melbourne and Greater Sydney urban regions after each exclusion step.

| SA1 regions | Greater Melbourne | Greater Sydney |
| --- | --- | --- |
| which were present in Pathzz data | 10378 | 11043 |
| for which URP, IEO, IER were available and mobility counts were non-zero | 10366 | 11026 |
| for which coverage was computable | 8809 | 9949 |

The Department of Transport of the Victorian state government collects pass-through counts of vehicles at intersections across Victoria at 15 minute intervals using the Sydney Coordinated Adaptive Traffic System (SCATS). The dataset is publicly available online [44]. We extracted the files from this dataset for the test and baseline periods chosen for our analysis of the Pathzz data. The counts are then temporally aggregated to obtain the mobility at each intersection for the test and baselines periods. In the SCATS dataset, intersections are identified by an identification number. A list of SCATS sites and a description of their location [45] was obtained from a Vicroads report [46] (Vicroads is a Victorian state government and private body responsible for managing roads in Victoria). The intersections in the above database are not geocoded (do not contain lat-lon coordinates) but do identify the road names associated with each intersection. To identify the appoximate latitude and longitude of each intersection, we use string matching to perform geocoding of intersections from OpenStreetMap (OSM). The string matching is not perfect, and a single location description may yield multiple coordinates from OSM due to different roads or streets having the same name or due to OSM having the same information in different layers. In such cases, we assign the mean of all matching coordinates to the corresponding SCATS site. In order to remove unreliable matches between SCATS sites and geocoded intersections, we compute the standard deviation of the OSM coordinates and remove those matches whose standard deviation is greater than the diagonal of Melbourne metropolitan area’s largest intersection: Princes Highway and Warrigal Road in Oakleigh, Victoria.

We then associated the geocoded SCATS sites with the DZN partitions containing their coordinates. This allows a pair-wise comparison between the passthrough data from Victoria DoT and the Pathzz data for the corresponding DZN area. For each DZN, we calculate a measure to assess changes in mobility as the log-transformed ratio of counts during test and baseline periods:

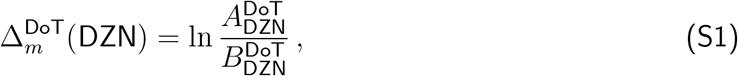

where

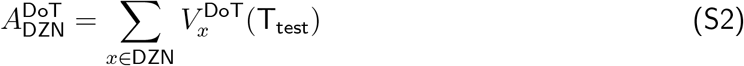

is the number of visits recorded for a destination zone, calculated by summing the visits, *V*_*x*_ at an intersection, *x*, over all intersections contained within that destination zone, during the test period *T*_test_ and

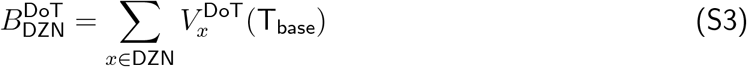

is the same measure for the corresponding pre-pandemic baseline duration *T*_base_.

For comparison, origin-destination counts from the Pathzz dataset were aggregated by destination, and the log transformed ratio of counts during test and baseline periods was computed:

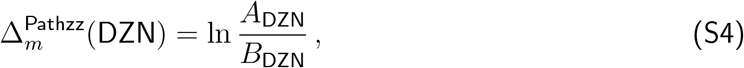

where

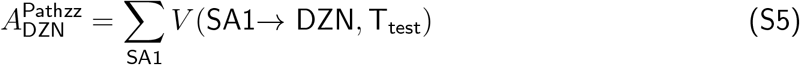

is the number of visits recorded for a destination zone, calculated by summing over the visits, V(SA1*→* DZN, from all non-intersecting SA1s during the test period *T*_test_ and

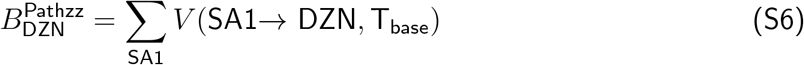

is the same measure for the corresponding pre-pandemic baseline duration *T*_base_.

We computed Spearman’s rank correlation between the following pairs of quantities: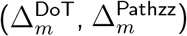, (*A*^DoT^, *A*^Pathzz^) and (*B*^DoT^, *B*^Pathzz^), after removing the outliers (values under 3 percentile and above 97 percentile). We found small yet positive correlations (*≈* 0.2) with significant p-values. All the lower confidence intervals (for 95%) were greater than 0.1. The data are shown as heatmaps in Figure S1 and results of Spearman correlation are summarised in Table S2.

##### 1. Classification of intersections from OpenStreetMap

OpenStreetMap (OSM) is an open-source, global geodatabase that includes detailed descriptions of the road networks of urban regions throughout the world. While the urban region of greater Melbourne is well-documented in the geodatabase, intersection points of roads are not classified directly by OpenStreetMap. To geolocate intersections identified in the SCATS database, we used the following process:

**FIG. S1.**
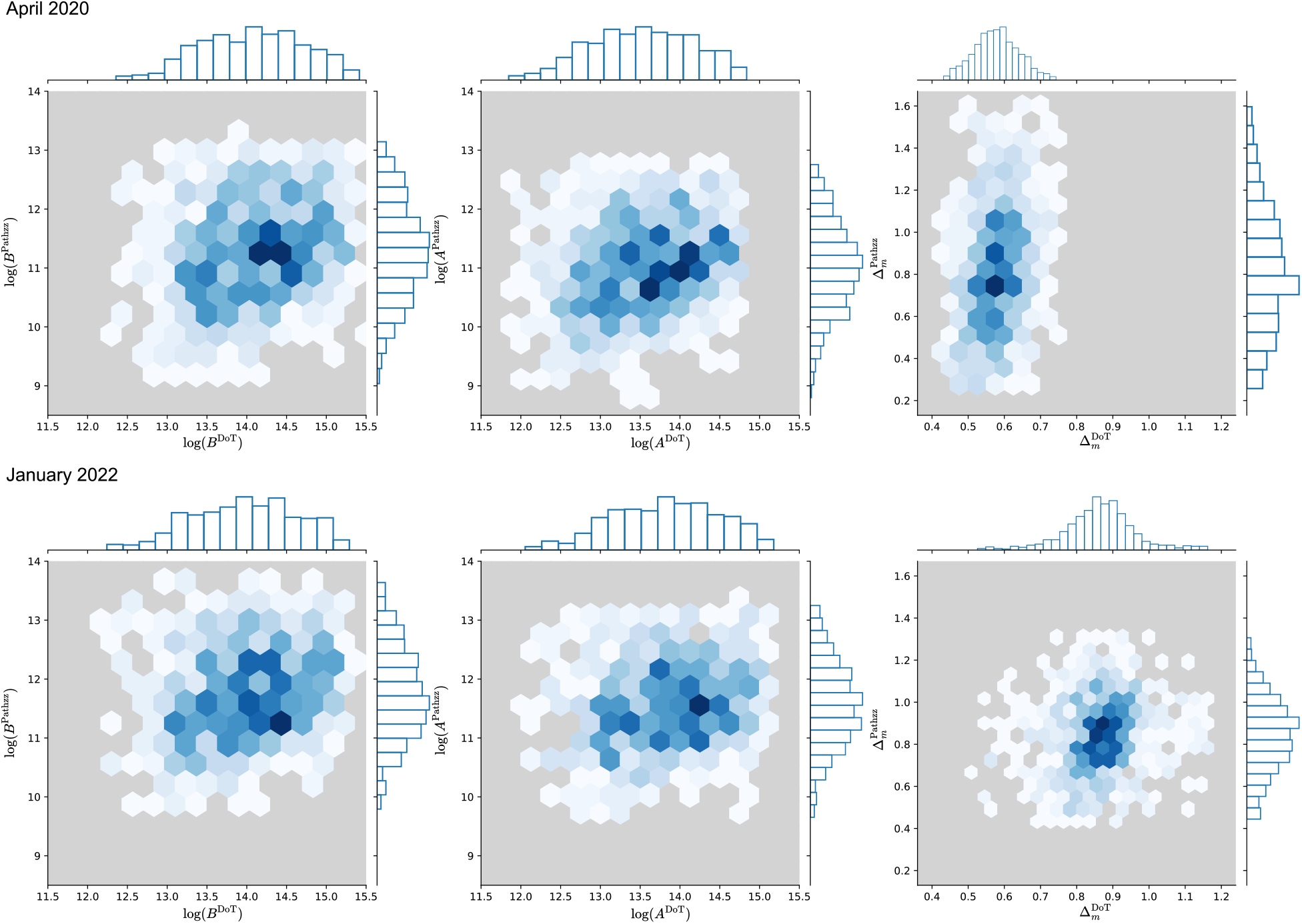
Heatmaps of mobility counts and measure of change in mobility from the two independent datasets (Pathzz and DoT).

- Identify roads as line objects provided by OpenStreetMap
- Identify intersections of roads as points joining the features of the intersecting linear road objects

Linear vector features representing roads were extracted from the OSM database using the Quick-OSM plugin for QGIS which allows a user to import OSM objects using the Overpass API. Objects are imported using key-value pairs from the OSM database. For this analysis, road objects were imported over the spatial extent of greater melbourne according to the key-value pairs listed in Table S3. To geolocate intersections, the intersection points of roads were identified as point features by using the Vector Overlay: Line Intersections tool in QGIS. The latitude and longitude of each point was then computed using the Vector Geometry: Add Geomery Attributes tool. The resulting set of intersection points were linked with the DZN partitions containing them for key values highway motorway OR motorway link OR motorway junction highway trunk OR primary highway secondary OR secondary link highway tertiary OR tertiary link highway residential validation with SCATS data as described above.

**TABLE S2.**
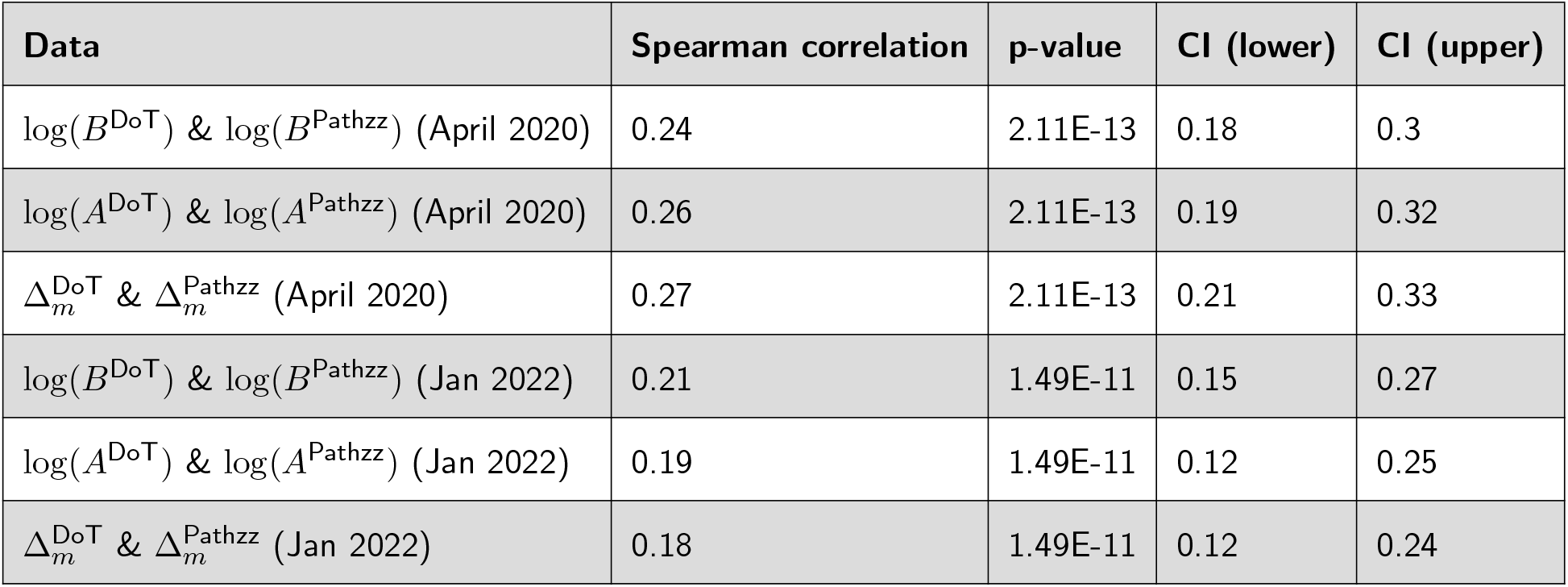
Spearman rank correlation (coefficient, p-value and 95% confidence intervals) between destination aggregated counts from the two datasets and the ratio of the counts (the test and its corresponding baseline period) from the two datasets.

| Data | Spearman correlation | p-value | CI (lower) | CI (upper) |
| --- | --- | --- | --- | --- |
| $\log(B^{\text{DoT}}) \& \log(B^{\text{Pathzz}})$ (April 2020) | 0.24 | 2.11E-13 | 0.18 | 0.3 |
| $\log(A^{\text{DoT}}) \& \log(A^{\text{Pathzz}})$ (April 2020) | 0.26 | 2.11E-13 | 0.19 | 0.32 |
| $\Delta_m^{\text{DoT}} \& \Delta_m^{\text{Pathzz}}$ (April 2020) | 0.27 | 2.11E-13 | 0.21 | 0.33 |
| $\log(B^{\text{DoT}}) \& \log(B^{\text{Pathzz}})$ (Jan 2022) | 0.21 | 1.49E-11 | 0.15 | 0.27 |
| $\log(A^{\text{DoT}}) \& \log(A^{\text{Pathzz}})$ (Jan 2022) | 0.19 | 1.49E-11 | 0.12 | 0.25 |
| $\Delta_m^{\text{DoT}} \& \Delta_m^{\text{Pathzz}}$ (Jan 2022) | 0.18 | 1.49E-11 | 0.12 | 0.24 |

**TABLE S3.**
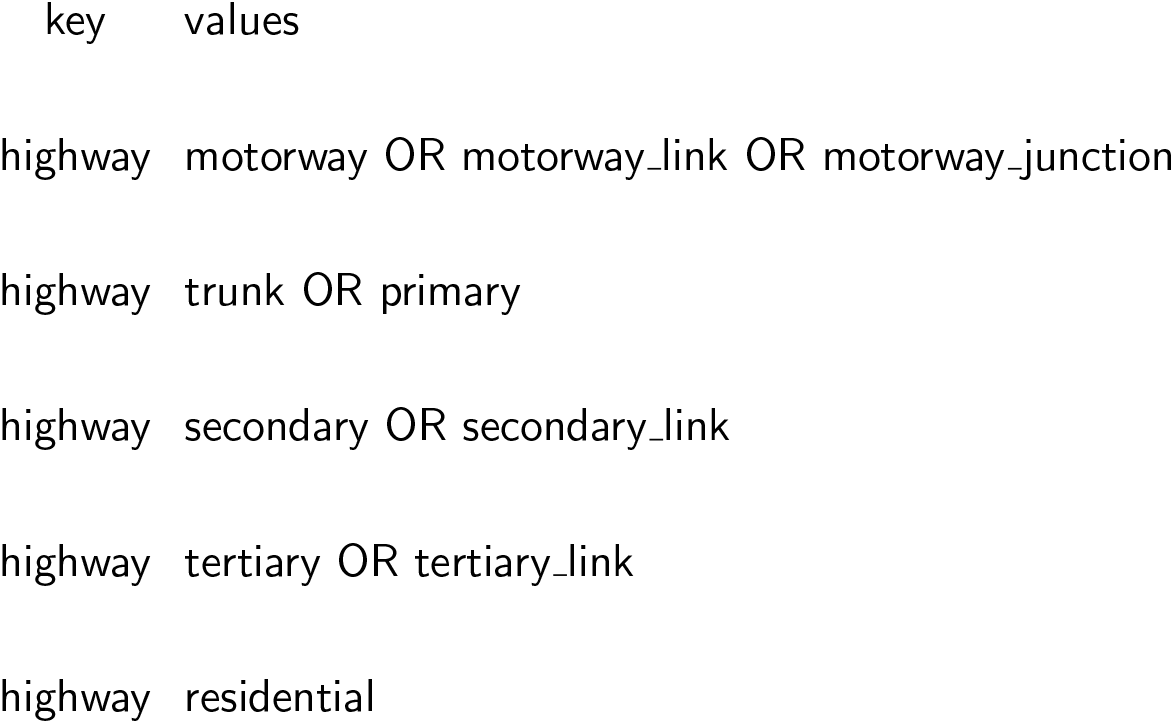
Key-value pairs used to extract features from OpenStreetMap for the purposes of intersection geocoding.

### S2. SUPPLEMENTAL DATA

**FIG. S2.**
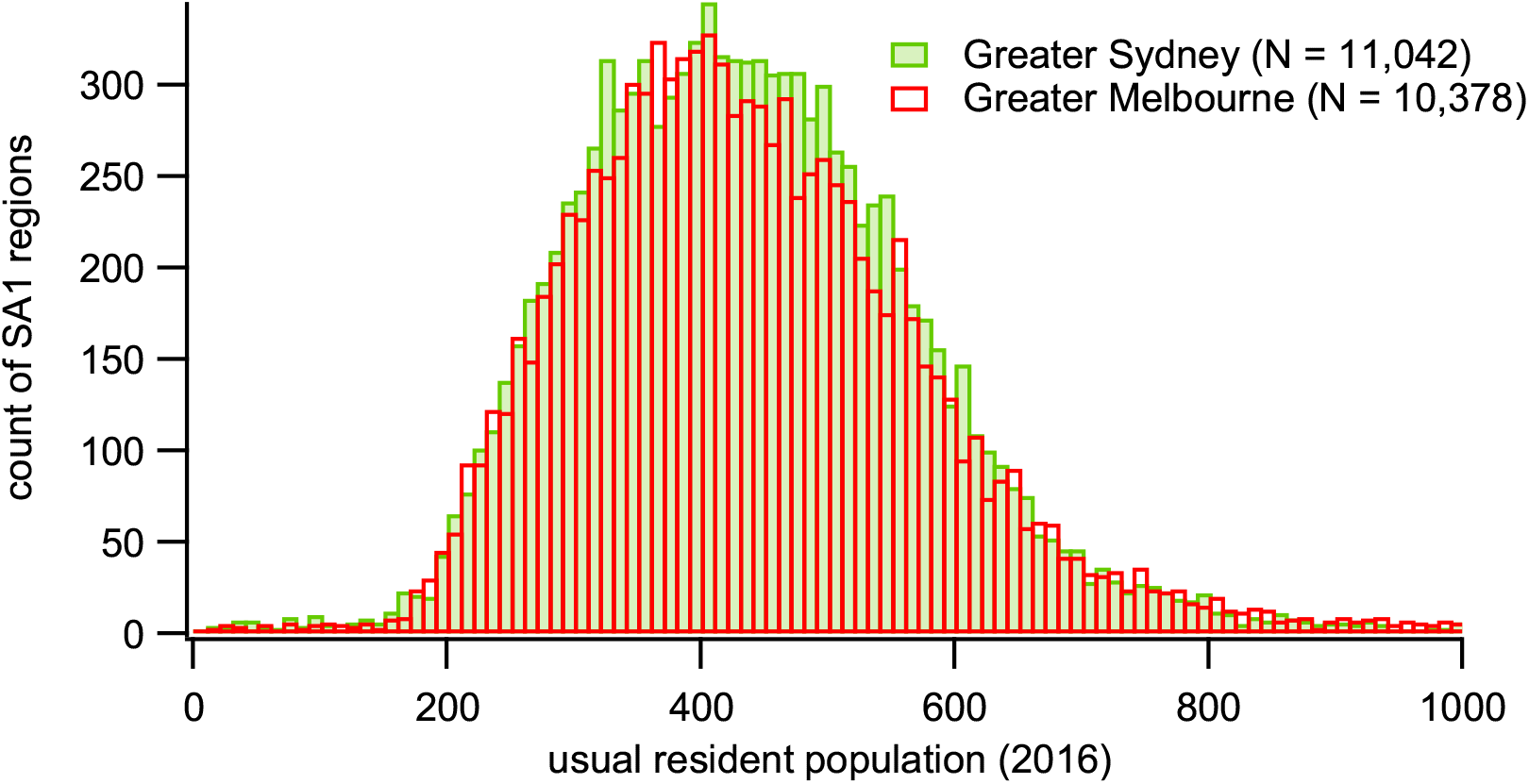
Distribution of usual resident population, as counted in the 2016 Australian Census, for the SA1 regions contained within the greater urban areas of Sydney and Melbourne.

**FIG. S3.**
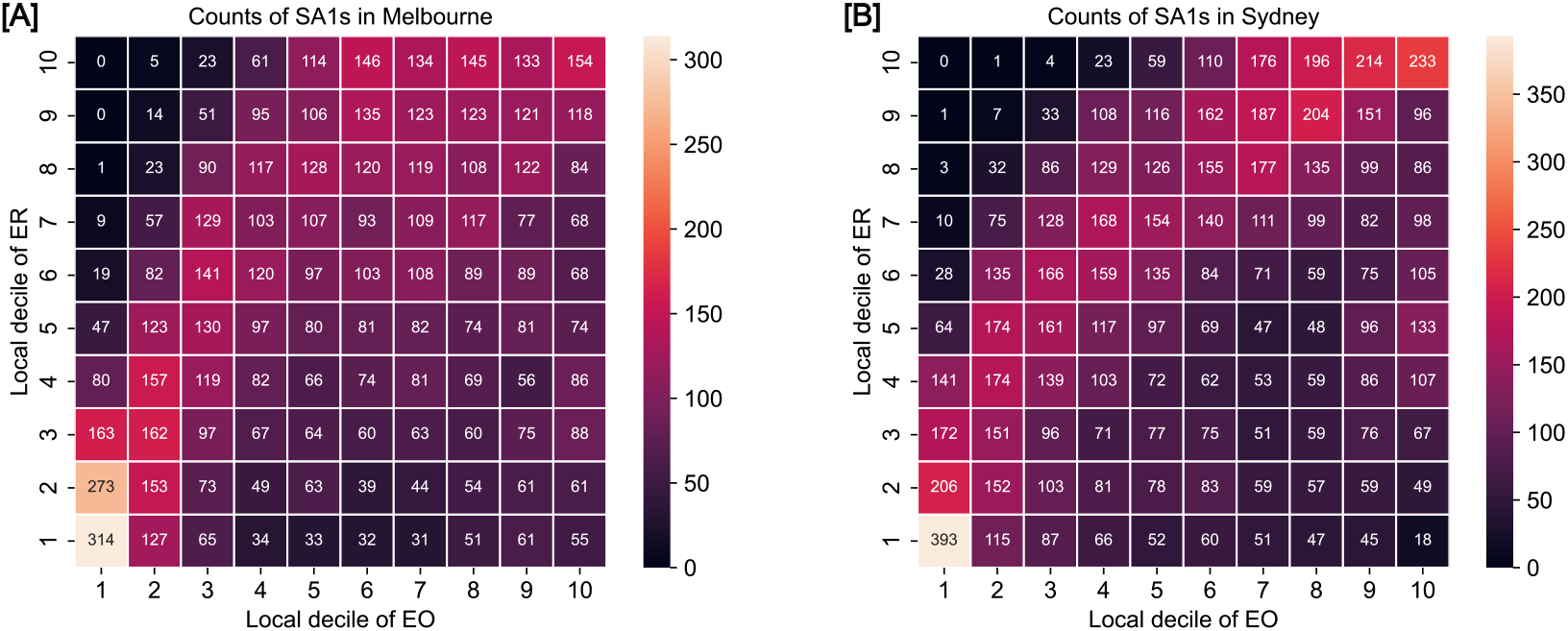
Number of of SA1 regions in each jointly-stratified ER-EO decile.

**FIG. S4.**
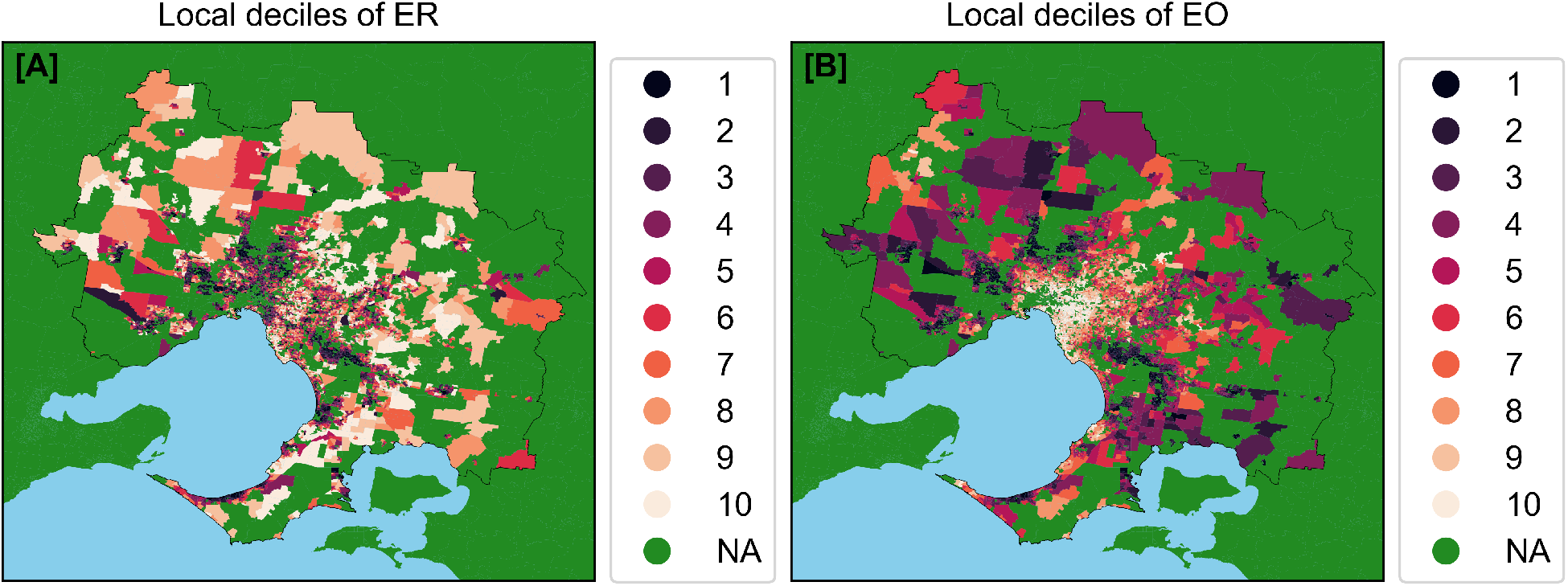
Map of Greater Melbourne showing the deciles of ER (A) and EO (B). Gray color (NA in the legend) indicates those regions which were excluded from the analysis because they were either outside of the Greater Melbourne region or because their coverage could not be computed. The black line shows the boundary of the Greater Melbourne urban region. Blue color indicates sea.

**FIG. S5.**
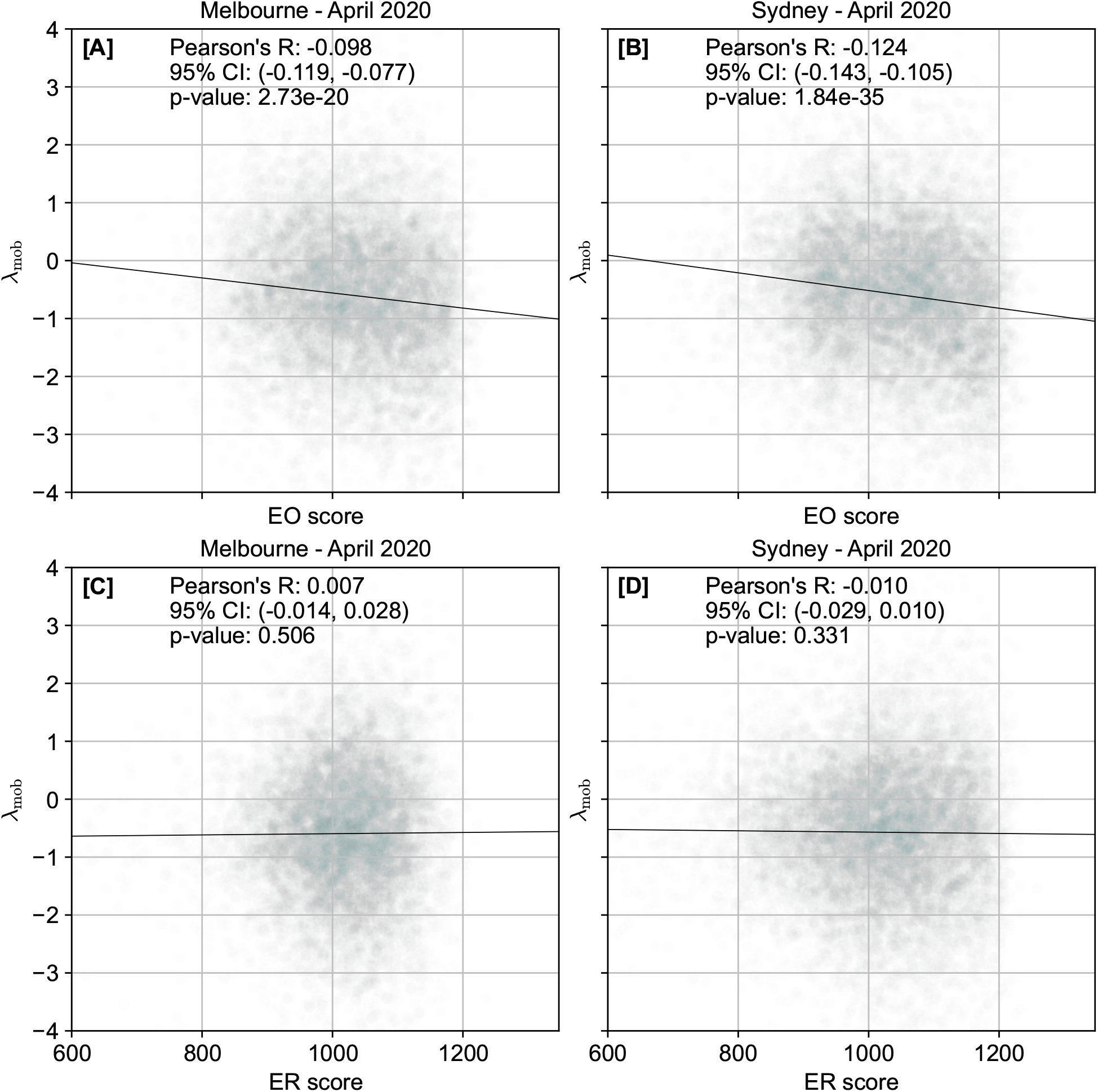
Scatter plot of *λ*_mob_ against various SEIFA scores in Melbourne and Sydney during April 2020. Pearson’s correlation coefficient is shown on the plots, along with the 95% confidence intervals. The black colored straight lines show the regression slope. Outliers are not shown in the plot for ease of visualisation, but are included in the correlation computation.

**FIG. S6.**
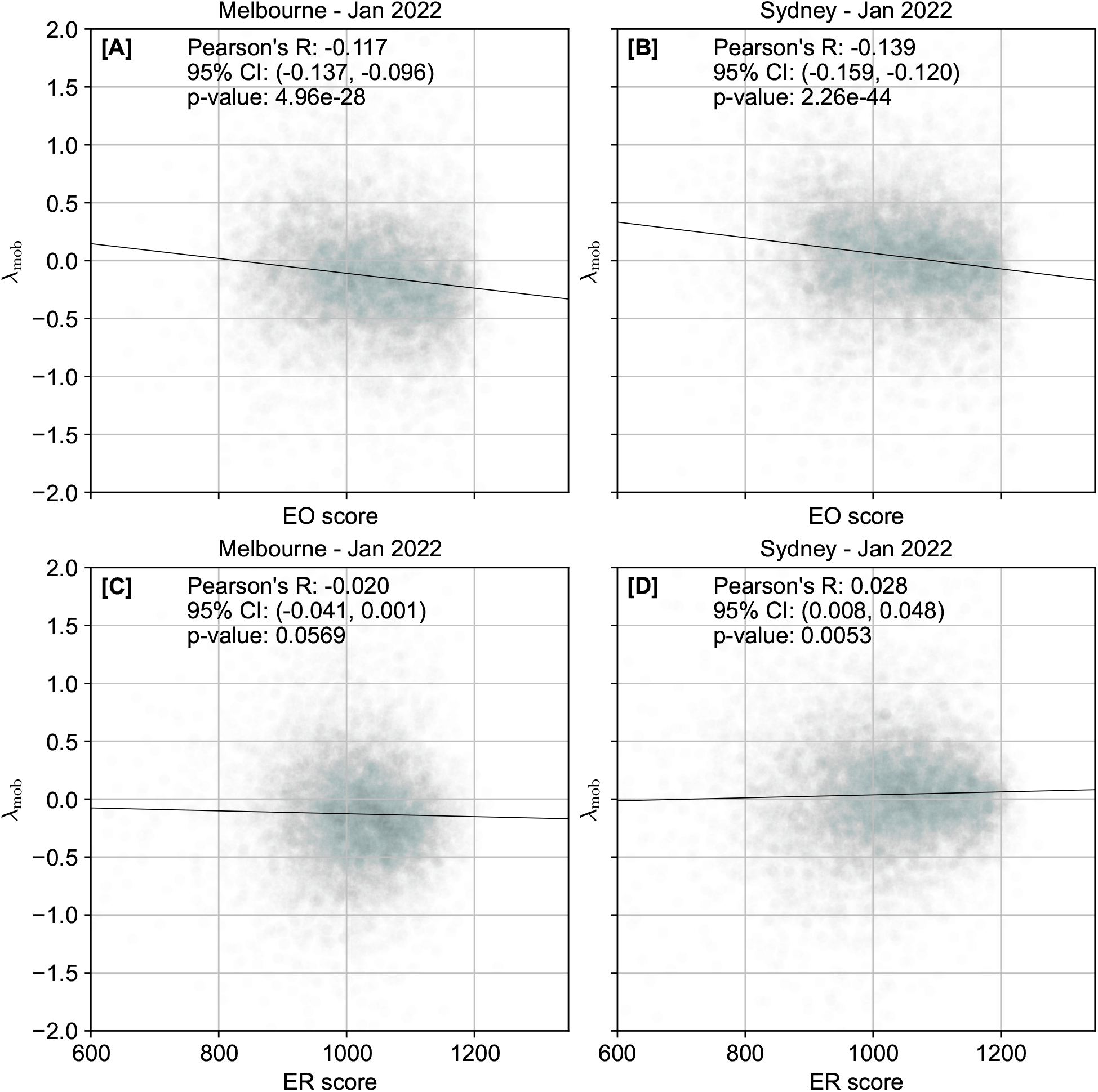
Scatter plot of *λ*_mob_ against various SEIFA scores in Melbourne and Sydney during January 2022. Pearson’s correlation coefficient is shown on the plots, along with the 95% confidence intervals. The black colored straight lines show the regression slope. Outliers are not shown in the plot for ease of visualisation, but are included in the correlation computation.

